# Intermittent theta burst stimulation for attention deficit hyperactivity disorder

**DOI:** 10.1101/2025.09.03.25335033

**Authors:** Brian C. Kavanaugh, Megan M. Vigne, Christopher Legere, Gian DePamphilis, Eric Tirrell, W. Luke Acuff, Joshua Brown, Rich Jones, Anthony Spirito, Linda L. Carpenter

**Affiliations:** E.P. Bradley Hospital; Brown University, Department of Psychiatry & Human Behavior; Butler Hospital TMS Clinic and Neuromodulation Research Facility; Providence Veteran’s Association Medical Center, Center for Neurorestoration and Neurotechnology; Brown University, Department of Neuroscience

**Keywords:** Theta burst stimulation, attention deficit hyperactivity disorder, working memory, anger, adolescence

## Abstract

**Objective**: Attention deficit hyperactivity disorder (ADHD) is the most diagnosed psychiatric disorder in childhood and often causes lifelong symptom burden. Transcranial magnetic stimulation (TMS) has been investigated in adult ADHD with encouraging findings, although work in pediatric samples remains limited and no ADHD studies have examined the utility of intermittent theta burst stimulation (iTBS). **Methods**: Twenty-nine adolescents with ADHD and working memory (WM) symptoms were randomized into a sham-controlled, counter-balanced, double-blind, crossover clinical trial of iTBS for adolescent ADHD. Participants completed ten active iTBS sessions (600 pulses per session) and ten sham iTBS sessions to the left dorsolateral prefrontal cortex (DLPFC) at 80% resting motor threshold. Primary outcome variables included safety, feasibility, and change in parent-reported ADHD and WM symptoms. Secondary outcomes consisted of parent and participant-reported affective symptom changes. **Results**: The protocol was feasible (82% completed all scheduled sessions) and safe (zero serious adverse events). A statistically significant improvement was seen in active versus sham iTBS in parent-reported overall ADHD symptoms, hyperactivity/impulsivity, working memory, anger, depressive symptoms, and anxiety symptoms. **Conclusions**: iTBS holds promise as a potential future treatment for ADHD, and effects achieved when targeting the left DLPFC may be most robust for transdiagnostic cognitive and affective symptoms. Increasing the number of iTBS sessions per day with accelerated protocols may maximize efficacy and feasibility for teens with ADHD and their parents. Clinicaltrials.gov Identifier: NCT05102864

## INTRODUCTION

Attention deficit hyperactivity disorder (ADHD) is a common neurodevelopmental disorder that affects approximately 10% of youth and is characterized by a persistent pattern of inattention and/or hyperactivity-impulsivity(1, 2). Ninety percent of children with ADHD continue to experience residual symptoms into young adulthood, and only 9% experience full recovery in adulthood(3). ADHD is a disorder of delayed cortical maturation particularly characterized by delayed peak thickness in prefrontal regions(4), and meta-analytic findings have shown reduced gray matter volume and task-related hypoactivation in multiple frontoparietal and limbic regions(5).

Non-invasive brain stimulation approaches to ADHD show tremendous promise. A recent meta-analysis found a medium effect size for rTMS to the dorsolateral prefrontal cortex (DLPFC) efficacy on overall ADHD symptoms(6). Six of the eight examined studies were completed with adults. In fact, only four ADHD studies have been done in children and/or adolescents (total n=106), including two sham-controlled trials (total n=52)(7–10). Utilizing a crossover design (n=9), Weaver and colleagues found no difference between active and sham rTMS in ADHD symptom change following 10 daily sessions of 10 Hz to the right DLPFC(9). However, Cao and colleagues(8) found ADHD symptom decrease (i.e., inattention, hyperactivity/impulsivity, and oppositionality) with active TMS (but not with sham TMS) after 30 daily sessions of 10 Hz to the right DLPFC (n=43).

In healthy adults, small-effect sized improvements in motor impulsivity are identified when applying rTMS to the left DLPFC, right inferior frontal gyrus, right frontal eye fields, and medial prefrontal cortex(11), and medium-effect sized improvements in temporal impulsivity are identified when targeting the left DLPFC with rTMS(11). Meta-analytic findings across neuropsychiatric disorders have shown that left DLPFC stimulation produces small effects on working memory, but no effect on attention(12). Another meta-analysis of left DLPFC rTMS in ADHD samples found a medium-sized effect on sustained attention(13). Preliminary work with rTMS targeting the right DLPFC(14) has also found beneficial effects on aspects of emotion regulation, and particularly anger in adults with PTSD(14). Similarly, in adults with MDD, irritability is significantly reduced following with intermittent theta burst stimulation (iTBS)(15) to the left DLPFC.

iTBS is particularly promising given its non-inferiority to 10 Hz rTMS for treating depression(16), shorter administration time, and feasibility as multiple sessions per day via accelerated iTBS protocols(17, 18). We conducted a trial of iTBS in adolescents with ADHD with a sham-controlled, double-blind, crossover design. We hypothesized that iTBS would be safe for participants and feasible for participants and their families, and that active iTBS would show superiority to sham in improving core ADHD symptoms, working memory, and emotional symptoms. We also conducted an open-label trial of twice-daily iTBS in a subsample to evaluate the viability of an accelerated iTBS schedule approach.

## METHODS

### Design

This was a randomized, double-blind, sham-controlled, crossover design clinical trial examining iTBS for adolescent ADHD (clinicaltrials.gov; NCT05102864). Local Institutional Review Board approval and an investigational device exemption (IDE) from the United States Food and Drug Administration (FDA) were obtained. Thirty-four adolescents (12-18 years old) with ADHD were enrolled from November 2021 to October 2024 (Figure 1). Primary <u>inclusion criteria</u> were: a) a clinical diagnosis of ADHD (confirmed with parent-reported symptoms on the Vanderbilt Assessment Scales), b) parent-reported working memory symptoms (i.e., t-score≥ 60 on the Behavior Rating Inventory of Executive Function– Second Edition [BRIEF-2]), c) full-scale intelligence quotient≥ 80 (based on Wechsler Abbreviated Scales of Intelligence, Second Edition [WASI-II]), and English fluency. <u>Exclusion criteria</u> included active mania, psychosis, substance use, current suicidal ideation, and standard TMS and MRI contraindications including history of seizures, participant or family diagnosis of epilepsy, significant current neurological disorder or intracranial pathology, prior brain injury, significant active medical condition, intracranial implanted metal devices, non-removeable metallic makeup or piercings, and chronic use of medications known to significantly decrease seizure threshold. Participants were not required to discontinue any clinical treatments, including psychostimulant medication, although they were asked to maintain consistency in medications throughout the trial.

**Figure 1.**
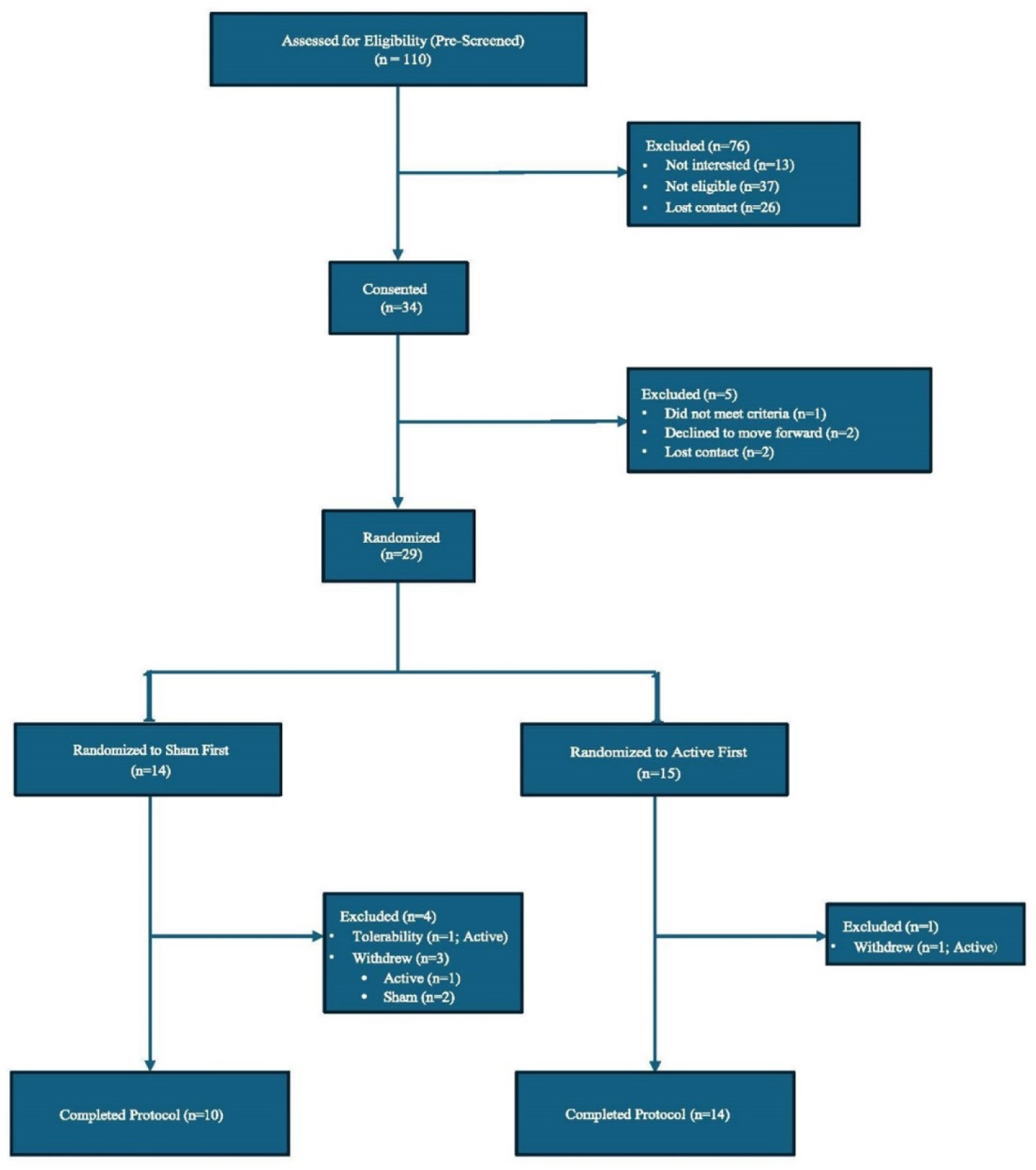
CONSORT flow diagram for a study of intermittent theta-burst stimulation (iTBS) for ADHD.

Participants completed two phases, with iTBS administered during the first 10 visits of each phase. Symptom status was gathered before the first iTBS session, immediately before the 6^th^ iTBS session, and ∼24 hours after the last session. A washout between phases was 6 weeks, except for one participant who waited four weeks due to sports season conflict.

### Treatment Intervention

10 iTBS sessions was selected based on prior iTBS studies available at the time of study design(19, 20) and prior TMS studies utilizing a crossover design to modulate neurocognitive processes(21, 22). We targeted the left DLPFC given the limited and mixed findings in ADHD targeting the right DLPFC(8, 9) and the more established left DLPFC findings associated with working memory-related processes(23).

Stimulation was delivered using a Nexstim NBT System 2 device with a figure-eight coil. Single pulses were used for mapping the left motor cortex for optional location and determining minimal threshold intensity for eliciting motor evoked potentials (MEPs ≥50 μV) in the contralateral abduces pollicus brevis muscle, with an integrated algorithm over at least 16 trials. The iTBS protocol was administered in 2-second trains of triplet 50 Hz bursts with 8-second rest intervals, for a total of 600 pulses/session, at 80% intensity relative to rMT. Each session lasted approximately three minutes.

### DLPFC Target

Structural MRI scans were completed prior to the first iTBS session. Stimulation was delivered to the left DLPFC target using individual participant MRI-based neuronavigation for coil placement during all sessions (Figure 2). Targeting was well-aligned across participants when examined in standard Montreal Neurological Institute (MNI) coordinates (Figure 2). The left DLPFC target site was identified based on anatomical landmarks per Mylius and colleagues(24) (See Supplement for further details); The mean E-field max for each session was recorded and averaged across sessions during statistical analyses to check for equal dosing across groups.

**Figure 2.**
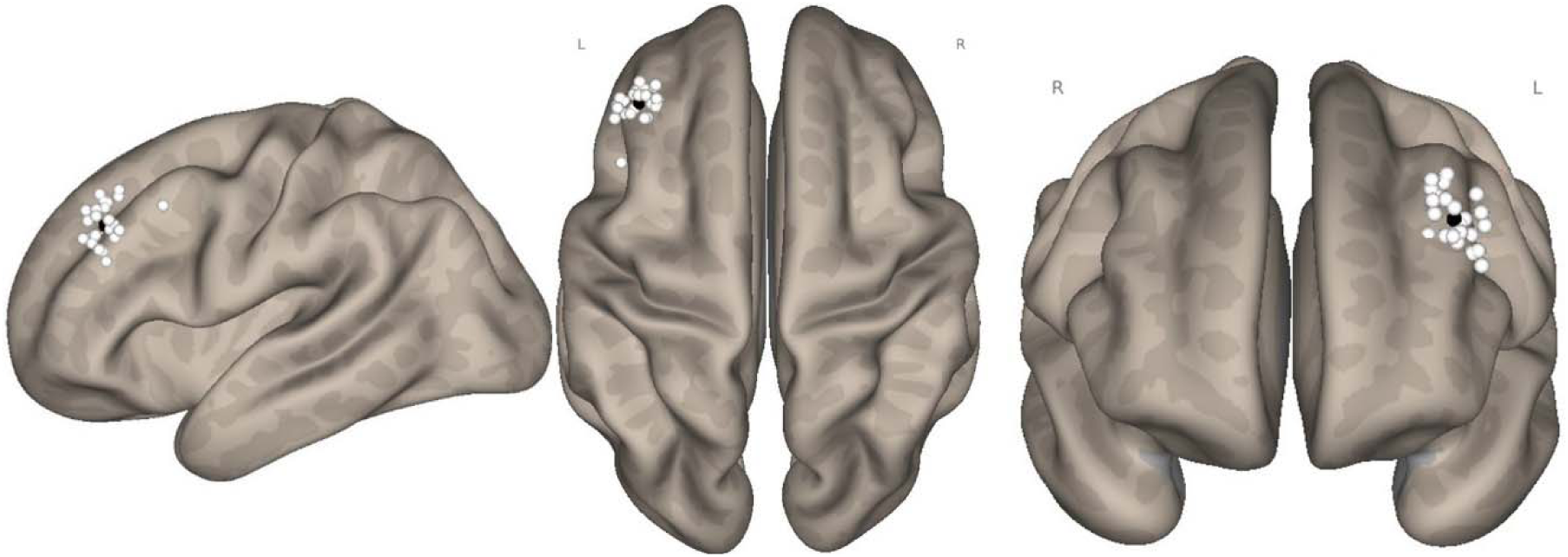
Target locations using the Mylius et al., 2013 method. **Legend**. White dots reflect individual participant stimulation target sites plotted on a standardized brain. Black dot reflects the group average MNI coordinates of participants in this study (−40, 36, 37).

### Randomization/Blinding

Participants were randomized 1:1 to receive active iTBS followed by sham iTBS (“Active First”) or sham iTBS followed by active iTBS (“Sham First”). Identical active and sham coils each had a colored label corresponding with their treatment condition. Each participant was assigned to either a “yellow” or “rainbow” condition. The principal investigator and research treatment team remained masked to the condition associated with each coil. Participants were asked to guess if they received “real” or “fake” TMS; most were asked after their first iTBS session, although the first five participants were asked to guess after their first full phase of iTBS (both approaches were analyzed).

### Clinical Variables

The primary efficacy outcome measures were parent-reported: a) National Institute for Children’s Health Quality Vanderbilt Assessment Scale for ADHD(25) (VAS; total score for items 1-18 [“Total ADHD Score”] to assess 9 core inattentive symptoms [“Inattentive Score”] and 9 core hyperactive/impulsive symptoms [“Hyperactive/Impulsive Score”] of ADHD to assess core ADHD symptoms, and b) Working Memory subscale of the Behavior Rating Inventory of

Executive Function – Second Edition (BRIEF-2)(26) to assess working memory symptoms. The secondary efficacy outcome measure was the Patient-Reported Outcomes Measurement Information System (PROMIS) participant and parent proxy forms(27) to assess anger, anxiety, depressive symptoms, positive affect, and stress-related symptoms. The PROMIS (parent and participant report) forms were added to the study protocol after the sixth participant, after several parents anecdotally reported that reduced anger and/or irritability was one of the most striking changes observed in their child following completion of the study. Intelligence quotient (IQ) and performance-based executive functions were assessed with the WASI-II (2-Subtest IQ)(28) and NIH Toolbox Cognition Battery(29), respectively (Table 1). Diagnostic status was assessed by parent report based on clinical history; a subset (n=20) completed the Mini International Neuropsychiatric Interview (MINI-KID)(30). Clinical and demographic variables were parent-reported at the enrollment session. Raw scores were utilized for all measures except IQ.

**Table 1.**
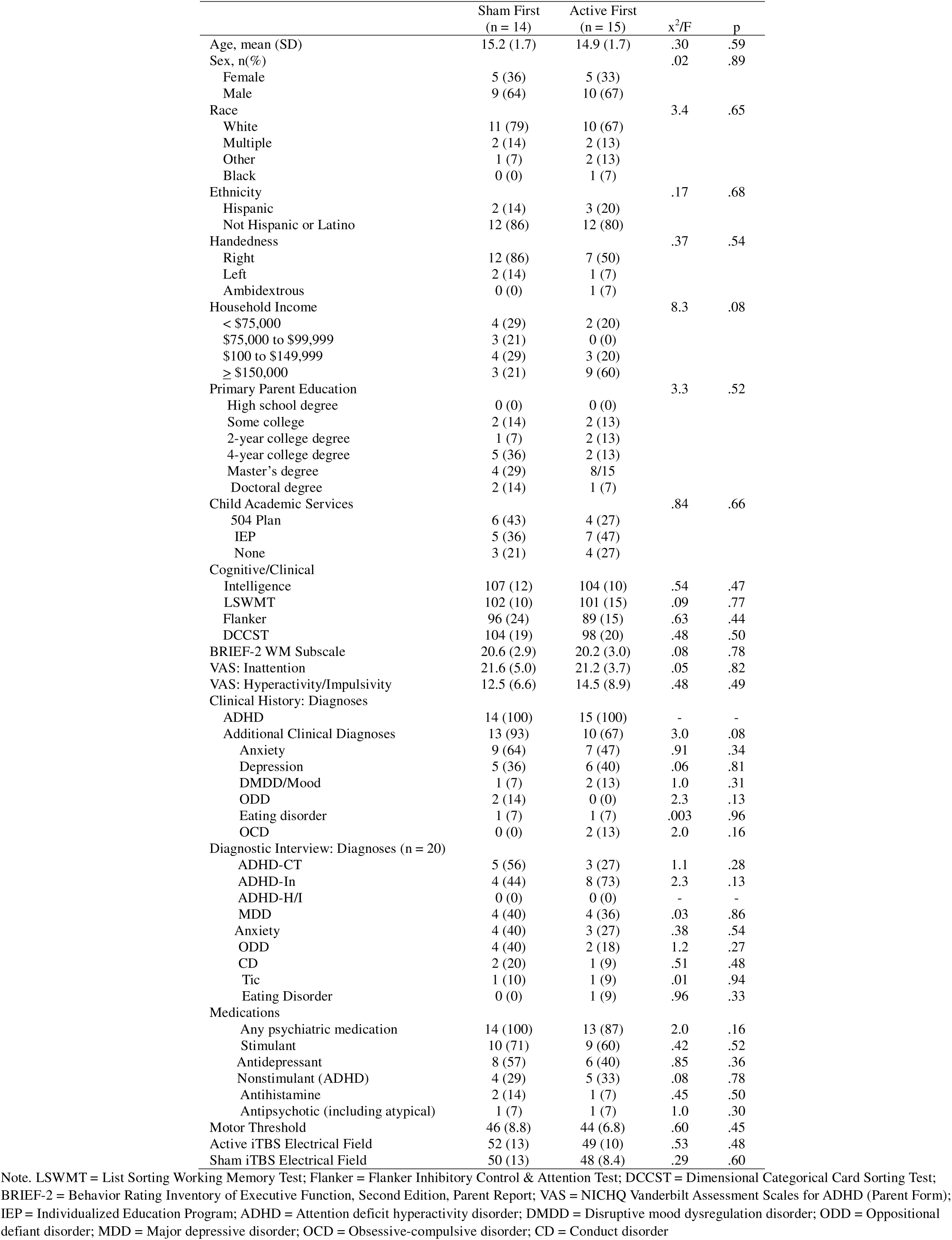
Demographic, clinical, and treatment characteristics of sample, grouped by order in which they received TMS.

### Statistics

Primary outcomes included: a) safety b) feasibility, and c) change in parent-reported ADHD and WM symptoms. Secondary outcomes consisted of parent- and participant-reported affective symptom changes. Primary outcomes for safety/feasibility are reported for all enrolled subjects who received at least one stimulation session (n = 29). Given the small sample size and preliminary nature of this study, efficacy analyses were conducted only with the sample of completers (n = 24). A series of two-way repeated measures analyses of covariance (ANCOVA) was conducted with treatment group (i.e., Sham First versus Active First; to account for order effects) and psychostimulant use (i.e., yes/no) as between-subject factors. Treatment condition (active or sham) by time (baseline, post-5 sessions, post-10 sessions) interaction was the primary outcome variable for all parent- and participant-report measures. In statistically significant models, pairwise comparisons of group differences after 5 and 10 sessions examined dose-response relationships. Estimated marginal means (EMMs) and standard errors (SE) are plotted in Figure 2, while raw score means and standard deviations (SD) are reported in Table 2. Effect size ([sham change – active change]/pooled SD) and % change were calculated based on the EMMs of the ANCOVAs. Response data generated by the BRIEF-2 and PROMIS forms were recoded so that the “never” option was set to zero prior to calculating change scores. To statistically examine time effects of the accelerated iTBS cohort, a series of two-sided t-tests (as the two cohorts had different treatment length) examined the symptom change between baseline (“Pre-iTBS”) and after 5 days (i.e., 10 iTBS sessions; “Post-10 iTBS”), and at one-week and one-month follow-up (after 10 days was not analyzed as only n=4 received 20 sessions). Statistical significance was set at p<.05.

**Table 2.** Clinical outcomes comparing active and sham intermittent theta burst stimulation.

|  | N | Pre-Session 1 | Sham<br>Post-Session 5 | Post-Session 10 | Pre-Session 1 | Active<br>Post-Session5 | Post-Session 10 | Condition x<br>Time | Cohen's d | % change<br>Sham | % change<br>Active |
| --- | --- | --- | --- | --- | --- | --- | --- | --- | --- | --- | --- |
| **Primary Outcomes** |  |  |  |  |  |  |  |  |  |  |  |
| BRIEF-2: Working Memory | 24 | 18.8 (3.8) | 17.6 (3.9) | 18.0 (3.8) | 19.4 (3.3) | 17.3 (3.7) | 16.8 (3.7) | F (2,19) = 4.5<br>**p = .03** | -.58 | 7.1 | 25 |
| VAS: Total ADHD Score | 24 | 20.6 (11.3) | 17.1 (11.5) | 16.7 (11.4) | 21.6 (11.8) | 17.0 (11.1) | 15.0 (9.4) | F (2,19) = 3.9<br>**p = .04** | -.48 | 13 | 36 |
| VAS: Inattention | 24 | 13.7 (6.8) | 11.8 (7.6) | 11.5 (7.0) | 14.0 (7.1) | 11.5 (7.2) | 10.1 (6.4) | F (2,19) = 2.3<br>p = .13 | -.41 | 13 | 31 |
| VAS: Hyperactivity/Impulsivity | 24 | 6.9 (5.9) | 5.3 (5.2) | 5.2 (5.5) | 7.7 (6.2) | 5.5 (5.3) | 4.8 (4.4) | F (2,19) = 6.3<br>**p = .01** | -.43 | 14 | 45 |
| **Secondary Outcomes (PROMIS Forms)** |  |  |  |  |  |  |  |  |  |  |  |
| Parent: Anger | 16 | 10.5 (4.0) | - | 10.1 (3.6) | 12.1 (4.4) | - | 9.3 (3.1) | F (1,12) = 8.9<br>**p = .01** | -.87 | -3.2 | 48 |
| Parent: Depression | 17 | 11.8 (4.8) | - | 10.8 (4.5) | 12.1 (5.8) | - | 9.7 (3.6) | F (1,13) = 10.1<br>**p = .01** | -.62 | 12 | 57 |
| Parent: Anxiety | 16 | 12.5 (4.2) | - | 12.4 (4.2) | 13.7 (5.1) | - | 13.3 (2.9) | F (1,12) = 24<br>**p < .001** | -.98 | -28 | 53 |
| Parent: Positive Affect | 17 | 12.4 (3.4) | - | 13.1 (3.3) | 12.9 (3.0) | - | 13.8 (3.2) | F (1,13) = .59<br>p = .46 | .18 | -10 | -18 |
| Parent: Stress | 17 | 10.4 (3.4) | - | 9.1 (3.8) | 10 (4.3) | - | 9.1 (2.6) | F (1,13) = 2.0<br>p = .18 | -.43 | 8 | 33 |
| Participant: Anger | 17 | 9.4 (3.5) | - | 9.0 (3.0) | 9.1 (3.4) | - | 9.0 (3.6) | F (1,13) = .76<br>p = .40 | -.18 | 5.8 | 17 |
| Participant: Depression | 17 | 15.0 (6.4) | - | 13.7 (6.0) | 14.5 (7.2) | - | 13.8 (7.2) | F (1,13) = .02<br>p = .89 | .02 | 13 | 10 |
| Participant: Anxiety | 16 | 13.8 (5.7) | - | 13.4 (6.4) | 13.3 (5.4) | - | 13.8 (6.8) | F (1,13) = .70<br>p = .42 | .13 | 6.8 | -7 |
| Participant: Positive Affect | 17 | 14.4 (3.8) | - | 14.8 (3.7) | 13.6 (3.2) | - | 13.8 (3.3) | F (1,13) = .01<br>p = .92 | .02 | -.9 | -2 |
| Participant: Stress | 17 | 8.8 (3.6) | - | 8.3 (4.0) | 9.4 (4.1) | - | 7.5 (3.5) | F (1,13) = 3.6<br>p = .08 | -.47 | -11 | 35 |

Note. Results reflect the repeated measures ANCOVA comparing active and sham iTBS on clinical outcomes. Raw scores are reported here. All other measures are parent-report unless participant-report is indicated. BRIEF-2 = Behavior Rating Inventory of Executive Function, Second Edition, Parent Report (Working Memory Subscale); VAS = NICHQ Vanderbilt Assessment Scales for ADHD (Parent Form; Inattention = Items 1-9 of VAS; Hyperactivity/Impulsivity = Items 10-18 of VAS; PROMIS = NIH Patient-Reported Outcomes Measurement Information System Forms (Parent and Participant-Report).

### Open-Label Pilot Study of iTBS Accelerated Schedule

After participating in the randomized clinical trial (RCT) described above, all participants in the RCT (regardless of study completion, outcomes) were invited to enter an open-label pilot study to examine the safety, feasibility, and preliminary efficacy of a twice-daily (“accelerated”) iTBS treatment schedule. This pilot study was initiated 2.5 years after opening the randomized crossover trial and enrolled 8 participants into one of two cohorts: n=4 received twice/day iTBS sessions (80% rMT), 1,800 pulses/session, over five consecutive weekdays, to the same left DLPFC target used in the RCT; n=4 received this same twice/daily protocol for ten days (20 total sessions; Supplemental Table 1). Symptoms were monitored in the same manner as the RCT: VAS, BRIEF-2 WM subscale, and PROMIS forms were administered pre-iTBS and post-iTBS, and at 1 week and 1 month follow-up. ADHD symptom response was defined as > 50% reduction in parent-reported symptoms from baseline score. ADHD symptom remission was defined as < 6 inattentive parent-reported symptoms occurring often to very often and < 6 hyperactive/impulsive parent-reported symptoms occurring often to very often (i.e., no longer meeting ADHD criteria).

## RESULTS

### Clinical Sample

As shown in Table 1, the sample is characterized as a predominantly male, white, and non-Hispanic cohort. Most of the sample had at least one clinical diagnosis in addition to ADHD (79%), particularly anxiety (55%) and depression (38%), most were receiving school-based services (76%), and most were prescribed psychiatric medication (93%), particularly psychostimulants (66%) and antidepressants (48%). Twenty-nine participants were randomized to a treatment group. There were no statistically significant differences between groups at baseline in clinical and demographic variables (Table 1). Symptom status at the start of each phase is provided in Supplemental Table 2.

### Safety

A total of 11 adverse events (AEs) were documented in the trial, across n=6 participants (over the total iTBS administered sessions of n=567), resulting in a 21% AE per participant rate. Ten of the 11 AEs occurred in the active iTBS phase. Nine of the AEs involved a mild headache or scalp discomfort that naturally subsided, one AE involved mild, transient “brain fog”, and one AE involved moderate, increased behavioral fatigue at school that was later attributed to mononucleosis. There were no serious AEs. One participant (3%) withdrew from the study due to poor tolerability (i.e., secondary to scalp discomfort) of stimulation (during active iTBS; i.e., 97% tolerability of sample).

### Feasibility

Twenty-four of the twenty-nine randomized participants (82%) completed both phases of the study and all study procedures (Figure 1). There was an average washout period between conditions of 8.6 +/- 2.9 weeks (range:4.1-15.4). Further, there was no correlation between length of washout period and symptom status at the start of phase 2 (all p>.05, including when just analyzing the group that received active iTBS in phase 1. Four participants withdrew from the study (3 of 4 while receiving sham iTBS) due to burdens associated with the requirement for daily treatment sessions at the clinic or due to other family or school demands. A total of 3 (10.3%) participants thus withdrew (for any reason) during sham iTBS (including n=1 who withdraw after completing sham iTBS phase) and 2 (6.9%) withdrew during active iTBS.

### Effectiveness of the Blind

When receiving active iTBS first, 50% of the participants guessed correctly (i.e., “real”), and although 82% of participants guessed correctly (“fake”) when sham iTBS (x^2^=2.7, p=0.10).

### Clinical Outcomes

Clinical outcomes were analyzed in the subsample that completed both active and sham phases of the trial (n=24). PROMIS measures (added during the trial) generated data in a subset of n=16. Among completers, a statistically significant difference in symptom change across time was identified between the treatment conditions (i.e., condition x time interaction) for parent-reported working memory symptoms (BRIEF-2 WM subscale; p=0.025; d=0.58; sham change: 7.1% vs active change: 25%), parent-reported overall ADHD symptoms (VAS Total Score; p=0.037; d=-0.48; 13% vs 36% change for sham and active, respectively), and parent-reported hyperactivity/impulsivity (VAS Hyperactivity/Impulsivity Score; p=0.006; d=-0.43; 15% vs 43% change), but not parent-reported inattention (VAS Inattention Score; p=0.18; d=-0.41; 10% vs 27% change; Table 2 & Figure 3.A). In pairwise condition comparisons in statistically significant models, condition differences were not significant after 5 sessions (p>.05) but were significant after 10 sessions (p<.05, Figure 3.A).

**Figure 3.**
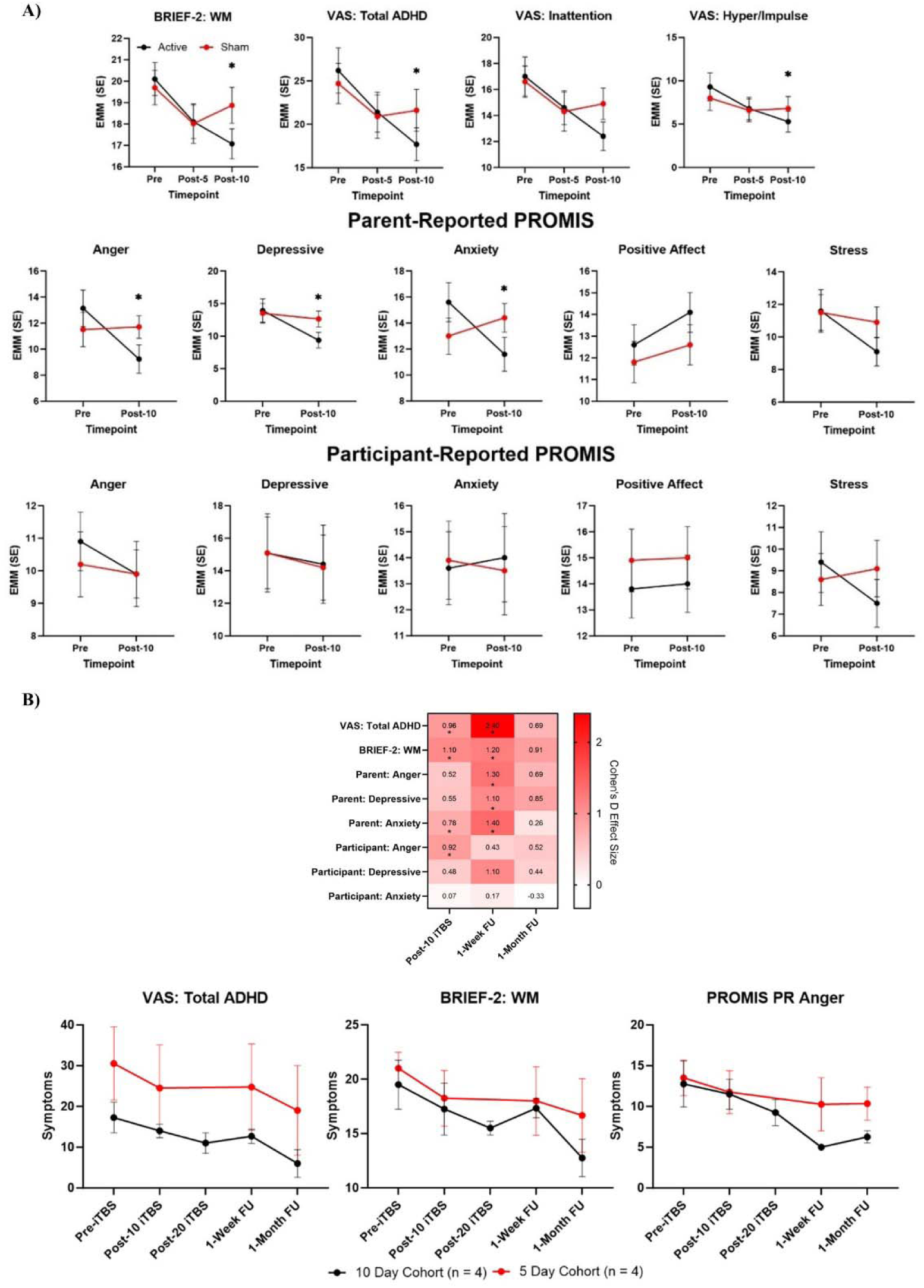
Clinical outcomes comparing active and sham intermittent theta burst stimulation and clinical outcomes of open-label twice-daily pilot study. **Legend**. A) Sham-controlled clinical trial. Results reflect the repeated measures ANCOVA comparing active and sham iTBS on clinical outcomes. Estimated marginal means from ANCOVA models are displayed here. * p < .05. Pairwise comparisons were only conducted on statistically significant overall ANCOVA models. B) Open-label twice-daily arm. Top: Effect size change in clinical symptoms in open-label pilot study of “accelerated” schedule (two sessions/day) iTBS. Two-sided t-tests examined change between Pre-iTBS and Post-10 iTBS, 1-Week Follow-Up, and 1-Month Follow-Up. *p < .05. Bottom: Raw scores displayed here at each time point for accelerated cohort (n = 8). Half the sample received 10 iTBS sessions (n = 4) and half the sample received 20 iTBS sessions (n = 4).

Among secondary outcome measures, a statistically significant condition x time difference was identified for change in parent-reported anger (PROMIS; p=0.01; d=-0.87; −3.2% vs 48% change), for change in parent-reported depressive symptoms (PROMIS; p=0.007; d=-0.62; 12% vs 57% change), and for change in parent-reported anxiety symptoms (PROMIS; p<.001; d=-0.98; −28% vs 53% change). No statistically significant group differences were identified on the five PROMIS participant-reported subscales or on the PROMIS parent-reported positive affect or stress subscales (Table 2 & Figure 3.A).

### Open Label Pilot Study of Accelerated Schedule

No adverse events (AEs) or early discontinuations occurred during twice daily iTBS in either the 5- or 10-day group (each n=4). One participant only tolerated stimulation intensity up to 45% MT for all visits. When examining the pooled cohorts (n=8) after 5 days of treatment, we observed categorical ADHD symptom response (i.e., >50% reduction from baseline VAS score) in one participant, and this response status persisted at one-month follow-up. Following 10 days of twice daily iTBS, three out of four (75%) participants experienced categorical ADHD symptom response and also ADHD symptom remission (i.e., <6 inattentive and <6 hyperactive/impulsive-type symptoms rated as occurring ‘often to very often’) that persisted at one-month follow-up.

In a series of t-tests examining pre-versus post-iTBS differences, statistically significant change after iTBS was observed in parent-reported total ADHD symptoms, working memory symptoms, anxiety symptoms, as well as participant-reported anger symptoms (n=8; all p<.05; Figure 3B). At one-week follow-up (n=8), statistically significant change was similarly observed in parent-rated total ADHD symptoms, working memory symptoms, anger symptoms, depressive symptoms, and anxiety symptoms. No statistically significant change from baseline persisted at one-month follow-up in the full cohort.

## DISCUSSION

This is the first study, to our knowledge, that has investigated iTBS for ADHD. Using a randomized, sham-controlled, double-blind, crossover design in adolescents with ADHD, we found that active iTBS to the left DLPFC was safe and feasible, and, consistent with our hypothesis, superior to sham in improving core ADHD clinical features, including symptoms related to working memory, hyperactivity/impulsivity, and anger.

Twenty-one percent of the randomized sample reported experiencing at least one non-serious, transient AE but no participant had a serious AE. All AEs were considered expected and typical for TMS, largely consistent with prior child/adolescent ADHD-TMS studies(7–10) and adult TMS studies. AEs included headache (22%), discomfort (11%), and pain (24%)(31). Twenty-four (83%) of the randomized sample completed the protocol (i.e., 17% discontinuation rate), which is consistent with prior child/adolescent ADHD-TMS clinical trials(7, 8).

Regarding preliminary efficacy, this is the third sham-controlled trial of rTMS efficacy in pediatric ADHD and the first to use iTBS. One of these prior studies found that 30 sessions of active rTMS (n=43; 10 Hz), but not sham rTMS, led to improvements in inattention, hyperactivity/impulsivity, and oppositionality(8), while the other study (n=9; 10 Hz) did not find an effect on ADHD symptoms after 10 sessions of active rTMS(9). A recent meta-analysis of rTMS in ADHD across the lifespan found a standardized mean difference of .45 for efficacy in reducing ADHD symptoms(6). This is highly consistent with findings reported here that also showed a medium effect size (d=.41-.48) for efficacy of iTBS on core ADHD symptoms. Most prior rTMS ADHD therapy trials have targeted the right DLPFC with scalp-based measurements (predominantly the “5 cm rule”), while the current study targeted the left DLPFC via individual MRI-based neuronavigation to an anatomically defined target. We utilized the working memory and the iTBS literature for selecting protocol parameters(19, 20, 23) given the TMS-ADHD literature was mixed at the time of study design(8, 9). The lack of similarity across studies makes it difficult to determine whether either of the targets (left versus right DLPFC) or targeting methods (5-cm, scalp-based versus MRI-based neuronavigation to anatomical target) are superior in optimizing efficacy for treatment of core ADHD symptoms. Related, we observed preliminary evidence of a dose-response relationship with the number of iTBS sessions in the RCT, in that there was no significant effect on any primary symptoms after 5 sessions, but the active versus sham iTBS differences emerged after 10 sessions for ADHD and WM symptoms. Future studies should test longer protocols than the current standard 10-session protocol.

The strongest effects in this study were not on core ADHD features, but rather on transdiagnostic and co-occurring symptoms, particularly parent-reported working memory (d=.58), anger (d=.87), depression (d=.62), and anxiety (d=.98). A published meta-analysis of data from adult trials showed a small effect of rTMS on performance-based working memory(12). Two prior studies have found that iTBS improved participant-reported anger symptoms (right DLPFC target) and participant-reported irritability symptoms (left DLPFC target) in adult PTSD(14) and MDD(15), respectively, with current findings extending prior work to show this effect is also observed in adolescent ADHD. While current effects on *parent-reported* anxiety and depression are consistent with the known effects of rTMS on adult anxiety(32) and depression(33), and even adolescent anxiety(34) and depression(35), no effects were found for *participant-reported* symptoms. This may reflect variable self-awareness and/or underreporting of symptoms phenomenon in ADHD(36). Alternatively, iTBS effects may not manifest in core emotional experiences but rather in cognitive control or behavioral manifestations of affect that are observed by parents. Additionally, alternative spatial targets should be examined for maximal targeting of core ADHD symptoms. While the left DLPFC may be optimal for working memory and mood, the right pre-SMA may be more optimal for inhibition and impulse control(11, 37), while the right the left superior frontal gyrus may be more optimal for core ADHD feature targeting(5).

Importantly, 14% of the sample withdrew from the study due to the demands of attending daily sessions. Although most of these families were receiving sham iTBS at the time of their withdrawal, we do think these findings are critical in treatment development. This protocol only administered 600 pulses per day (i.e., 3 minutes). Given the rapidly growing literature on safety and efficacy of protocols that administer numerous iTBS sessions per day(18), we conducted an open-label trial of twice-daily iTBS (3,600 pulses per day) for 5 or 10 days to previously enrolled participants in the clinical trial. In this small, sample of teens not naïve to iTBS, we found that twice-daily iTBS was safe (i.e., no AEs), feasible (no withdrawals), tolerated (i.e., 88% of the sample tolerated 80% MT), and effective for acutely improving parent-rated ADHD, working memory, and mood symptoms. In this small sample that received 36,000 pulses (20 total sessions), we observed a 75% response and remission rate of ADHD that persisted for at least one month (n= 3/4). Accelerated iTBS will likely show improved feasibility and comparable efficacy to traditional once per day iTBS for ADHD, although further work with large samples, sham-controlled trials, and treatment-naïve participants is needed.

There are several limitations to this study. First, the study had a modest sample size and a crossover design, so effect sizes may be an overestimation of true effects. Second, the DLPFC targeting approach used in this study has not previously been utilized in ADHD research, so an optimal or ideal cortical target for ADHD remains unknown. Third, consistent with other clinical trials in ADHD samples, self-report measures of ADHD were not collected from participants in this study, so results are dependent on parent observations and not internal experiences related to ADHD. Fourth, as this was a relatively short iTBS course (total 10 sessions), the optimal dose of iTBS for ADHD remains unknown. Finally, no long-term follow-up data was collected in the primary trial, so it remains unknown whether symptom improvement will persist.

## Data Availability

Data are available upon request

## Declaration of Interests

LLC has received support (through contracts with Butler Hospital) from Neuronetics, Affect Neuro, Janssen, Neurolief, Nexstim, and Biosynapse. She has received consulting income from Neuronetics, Janssen, Sage Therapeutics, Otsuka, Neurolief, and Magnus Medical. JCB receives financial support as an editorial stipend from Elsevier. BK, CL, GD, WA, ET, AS, MV, and RJ have no conflicts of interest. BK is supported by K23MH129853 and P20GM130452.

## Acknowledgements

This trial was funded by the NARSAD Brain & Behavior Research Foundation and the Carol Peterson Foundation.

## Supplemental Section

### Targeting

Mylius et al method uses anatomical landmarks on participant MRI to identify a left DLPFC target location corresponding with the BA46/BA9 junction in the Talairach atlas. It is identified as the location on the left middle frontal gyrus corresponding with the center of a line separating the anterior and middle thirds of the gyrus, and verified by anatomical placement on sagittal, coronal, and axial planes.

The Nexstim NBT System 2 software uses structural MRI data to generate an accurate 3D model of a person’s head and brain structures, with co-registration of landmarks on their head with landmarks on their structural MRI to allow tracking of the TMS coil position with respect to the underlying cortex. All participants wore a neoprene EEG cap (64 electrodes) during resting Motor Threshold (rMT) and iTBS treatment procedures for consistency across sessions (as some visits involved EEG recording). The Nexstim device models the induced E-field maximum (V/m) value on the cortical surface based on the individual participant’s anatomical MRI data and the physics of the coil.

**Supplemental Table 1.** Basic characteristics of open-label cohort.

|  | RCT Outcome | Days Between RCT and Open-Label | Open-Label Cohort |
| --- | --- | --- | --- |
| 1 | Completed | 676 days | 5-Day |
| 2 | Completed | 475 days | 5-Day |
| 3 | Completed | 92 days | 5-Day |
| 4 | Withdrew after randomization | 75 days | 5-Day |
| 5 | Completed | 20 days | 10-Day |
| 6 | Completed | 284 days | 10-Day |
| 7 | Withdrew prior to randomization | N/A | 10-Day |
| 8 | Withdrew prior to randomization | N/A | 10-Day |

**Supplemental Table 2.** Pre-iTBS primary clinical symptom status for each phase/coil.

| Outcome Variable | Sham First | Active First | F | p |
| --- | --- | --- | --- | --- |
| BRIEF-2 WM: Pre-Sham | 20.1 (3.1) | 17.5 (4.1) | 3.7 | .06 |
| BRIEF-2 WM: Pre-Active | 20.1 (2.5) | 18.8 (3.6) | 1.1 | .32 |
| BRIEF-2 WM: Pre-Phase 1 | 20.1 (3.1) | 18.8 (3.6) | 1.2 | .29 |
| BRIEF-2 WM: Pre-Phase 2 | 20.1 (2.5) | 17.5 (4.1) | 3.4 | .08 |
| VAS Total ADHD: Pre-Sham | 26 (14.4) | 16.9 (9.3) | 3.9 | .06 |
| VAS Total ADHD: Pre-Active | 23.8 (12.2) | 19.8 (11.4) | .74 | .40 |
| VAS Total ADHD: Pre-Phase 1 | 26 (14.4) | 19.8 (11.4) | 1.7 | .21 |
| VAS Total ADHD: Pre-Phase 2 | 23.8 (12.2) | 16.9 (9.3) | 2.6 | .12 |
| VAS Inattention: Pre-Sham | 16.5 (8.0) | 11.0 (5.3) | 4.46 | .04 |
| VAS Inattention: Pre-Active | 16 (6.7) | 12.7 (7.4) | 1.4 | .25 |
| VAS Inattention: Pre-Phase 1 | 16.5 (8.0) | 12.7 (7.4) | 1.8 | .19 |
| VAS Inattention: Pre-Phase 2 | 16 (6.7) | 11 (5.3) | 4.4 | .05 |
| VAS Hyperactivity/Impulsivity: Pre-Sham | 9.5 (7.9) | 5.9 (5.2) | 2.0 | .17 |
| VAS Hyperactivity/Impulsivity: Pre-Active | 7.8 (7.2) | 7.1 (5.4) | .08 | .78 |
| VAS Hyperactivity/Impulsivity: Pre-Phase 1 | 9.5 (7.9) | 7.1 (5.4) | .89 | .35 |
| VAS Hyperactivity/Impulsivity: Pre-Phase 2 | 7.8 (7.2) | 5.9 (5.2) | .58 | .45 |

Note. The group that received active iTBS in phase 1 had somewhat lower symptoms (none were $p < .05$ ) at the start of phase 2 than the group that received sham iTBS in phase 1. This ordering effect was not unexpected, which is why group status was loaded as a between-subjects factor in primary ANCOVAs.

## Notes

### Competing Interest Statement

The authors have declared no competing interest.

### Clinical Trial

NCT05102864

### Author Declarations

IRB approval and participant consent were obtained. Brown University Health IRB 410821

